# Neural basis of successful DBS for OCD after failed capsulotomy

**DOI:** 10.64898/2026.06.08.26355178

**Authors:** Melissa A. Ryan, Reem El Jammal, Sarah S. Soubra, Danika Paulo, Jonathan H. Bentley, Thomas A. Hamre, Nisha Giridharan, Hideo Suzuki, Nora Vanegas Arroyave, Eric A. Storch, Garrett P. Banks, Wayne K. Goodman, Nicole R. Provenza, Sameer A. Sheth, Sarah R. Heilbronner

**Affiliations:** Department of Neurosurgery, Baylor College of Medicine, Houston, TX USA 77030; Department of Neurology, Baylor College of Medicine, Houston, TX, USA 77030; Menninger Department of Psychiatry and Behavioral Sciences, Baylor College of Medicine, Houston, TX USA 77030; Rice Department of Electrical and Computer Engineering and Center for Neuroengineering, Rice University, Houston, TX USA; Neuroengineering, Rice University, Houston, TX USA Rice Department of Bioengineering, Rice University, Houston, TX USA; Rice Neuroengineering Initiative, Rice University, Houston, TX USA

**Keywords:** obsessive-compulsive disorder, capsulotomy, deep brain stimulation, connectivity, diffusion tractography, prefrontal cortex

## Abstract

**Background:** Obsessive-compulsive disorder (OCD) is characterized by disturbing thoughts (obsessions) that initiate anxiety-reducing thoughts or behaviors (compulsions). For patients with treatment-resistant OCD (tr-OCD), neuromodulation techniques, like capsulotomy (a lesion in the anterior limb of the internal capsule) and deep brain stimulation (DBS), have emerged as interventions that likely regulate connectivity between the prefrontal cortex (PFC) and subcortical targets. Three patients (Cap-DBS1-3) underwent a failed capsulotomy followed by successful DBS. Here, we aimed to understand the brain connections disrupted by failed capsulotomy vs modulated by successful DBS.

**Methods:** We used diffusion-weighted magnetic resonance imaging (dMRI) tractography in a control cohort with tr-OCD (n=12) and in two of the Cap-DBS patients themselves to determine connectivity profiles of the capsulotomy, volume of tissue activated (VTA), and potentially necessary tracts (VTA minus capsulotomy tracts). We used whole-brain, PFC-focused, and subcortically-focused tractography algorithms to fully explore the space of possible connections.

**Results:** Capsulotomy regions-of-interest (ROIs) connected with a variety of PFC and subcortical regions. VTA ROIs and potentially necessary tracts had limited and inconsistent PFC connectivity but substantial subcortical connectivity. While correlated to the average OCD connectome (r = 0.214, 95% CI [0.177, 0.251]; r = 0.756, 95% CI [0.739, 0.772]), the Cap-DBS connectomes had many edges that were stronger (z-score > 3).

**Conclusions:** The connectivity profile of potentially necessary tracts for successful DBS treatment after failed capsulotomy revealed a surprising proportion of subcortical regions and inconsistent PFC involvement, highlighting an often-ignored set of connections that may be critical to effective DBS.

**Key Messages:** *What is already known on this topic:* OCD has been conceptualized as a disorder of cortico-striato-thalamo-cortical (CSTC) circuits. Treatment targets have largely involved connections to the PFC.

*What this study adds:* Our study shows that potentially necessary tracts for DBS treatment after failed capsulotomy involve connections to subcortical regions and not PFC.

*How this study might affect research, practice, or policy:* This evidence of primarily subcortical involvement in treatment should encourage the use of more detailed subcortical atlases in neuromodulation circuit research and suggests that local circuit remodeling may be a mechanism of neuromodulatory OCD treatment.

## 1. Introduction

Obsessive-compulsive disorder (OCD) is a neuropsychiatric disorder that involves intrusive thoughts (obsessions) and behavioral or cognitive attempts to mitigate the distress induced by obsessions (compulsions). OCD has a prevalence of 1-3% in the United States ^1,2^ and ranges in severity. Many patients (40-60%) see symptom remittance from first line treatments, including psychopharmacology and cognitive-behavioral therapy ^3,4^. Some of the remaining patients, after not finding relief from multiple treatment attempts, are considered to have treatment resistant OCD (tr-OCD). In such cases, more invasive treatment approaches can be considered, including lesions (capsulotomies) and deep brain stimulation (DBS) of the ventral capsule/ventral striatum (VC/VS) ^5–8^. These treatments target white matter bundles to disrupt the neural circuitry involved in OCD pathology.

OCD is best characterized as a network disorder involving disparate brain regions that are interconnected in complex ways ^9–13^. Advances in functional and structural imaging have resulted in strong evidence for the involvement of the prefrontal cortex (PFC) and basal ganglia in the etiology of OCD ^14–18^. DBS and capsulotomy involve stimulation or lesions to PFC and basal ganglia pathways, although not exclusively. The regions or nuclei most impacted depend on both individualized neuroanatomy and the precise target chosen.

Although capsulotomies and DBS have comparable efficacy ^19,20^, they now appear to be in somewhat different locations. Early DBS VC/VS targets were based on the location of a typical capsulotomy ^21^; over time, DBS lead placement has trended posteriorly and ventrally relative to the initial targets ^22–24^. Still, given their proximity, there is a strong possibility that shared fibers between a typical capsulotomy lesion and a DBS volume of tissue activated (VTA) contribute to therapeutic outcome.

Regardless of precise placement, the brain structures and fibers in the vicinity of the VC/VS target are complex and not fully understood. The VC itself is the ventral part of the anterior limb of the internal capsule (ALIC), a large white matter bundle that bisects the caudate and the putamen/globus pallidus and carries fibers connecting the PFC to the thalamus, subthalamic nucleus, and brainstem ^25^. The VS is the limbic portion of the striatum, receiving inputs from ventral and medial regions of the PFC as well as the amygdala and hippocampus ^26,27^. While more anteriorly placed VTAs will intersect the VS, more posteriorly placed ones are more likely to intersect the bed nucleus of the stria terminalis (BNST), a small subcortical structure that is part of the extended amygdala ^28^. Laterally placed VTAs are situated near the globus pallidus externus. Activation of the fibers interconnecting these structures may also play a role in VC/VS therapeutic efficacy.

Previously, we reported on two patients who underwent successful DBS for OCD treatment after a failed capsulotomy ^29^. Because DBS and capsulotomy are thought to target many shared circuits, the idea that one would be successful following the failure of the other seems counterintuitive. However, in these cases, DBS leads were localized outside of the lesion site (Figure 1A), potentially implicating a different set of fibers.

**Figure 1:**
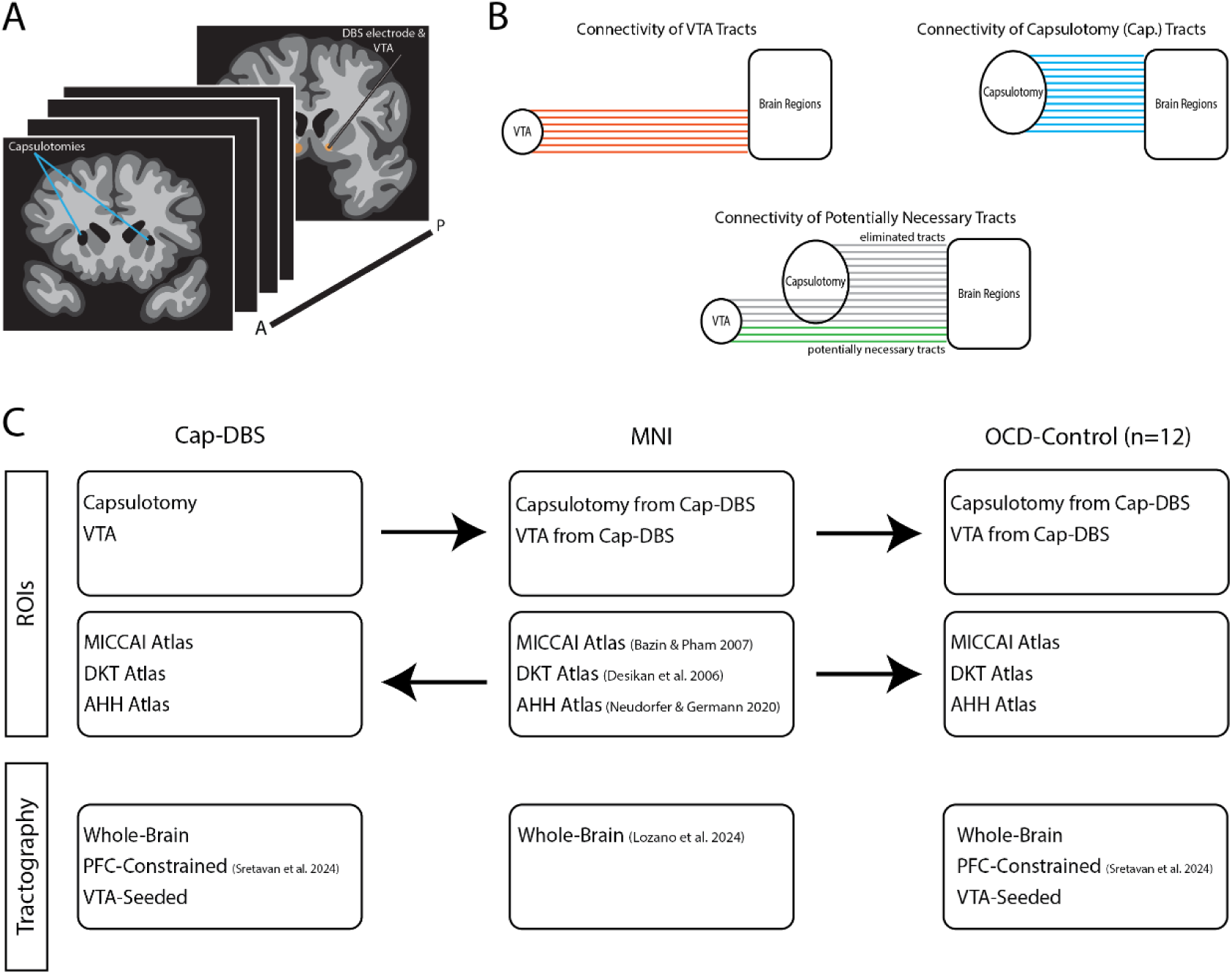
Analysis approach to investigate connectivity of VTA ROIs from Cap-DBS subjects. **A)** Illustration of capsulotomy and DBS VTAs in a single subject. Capsulotomies are more anterior and dorsal than VTAs. **B)** Schematics of tractography filtering performed to identify connectivity of Cap-DBS subjects’ capsulotomy and VTA ROIs. **C)** Schematic of the ROIs, atlases, and tractography datasets used in this study. *Abbreviations:* DBS, deep brain stimulation; VTA, volume of tissue activated; Cap, capsulotomy; ROIs, regions of interest; PFC, prefrontal cortex; MICCAI, Medical Image Computing and Computer-Assisted Intervention; DKT, Desikan-Killiany-Tourville; AHH, Atlas of the Human Hypothalamus

Here, we took a multi-pronged approach to understanding the circuits that enabled successful DBS following failed capsulotomy (Figure 1C). Because of the small number of patients treated with both capsulotomy and DBS, in addition to patient-specific diffusion tractography, we asked about the connectivity of the DBS VTA and the capsulotomy lesion in a tr-OCD control cohort. We were particularly interested in those fibers that are likely necessary for successful neuromodulation treatment: streamlines that were not impacted by the lesion but were within the VTA.

## 2. Methods

### Participants

We retrospectively analyzed data from three male patients (“Cap-DBS1-3”) who underwent a capsulotomy prior to undergoing DBS. Clinical response was defined by a ≥ 35% reduction in Y-BOCS score from the baseline assessment preceding DBS activation^30^. Details of response timelines are provided in Supplementary Methods. The “OCD-Control” cohort was composed of 12 patients with tr-OCD (50% male). Individuals in this cohort went on to be treated with DBS (see ^31^).

### MRI Acquisition

High-resolution magnetic resonance imaging (MRI) scans were obtained on a 3 Tesla Siemens MAGNETOM Prisma Fit scanner (Siemens Healthineers, Erlangen, Germany) at the Baylor College of Medicine (BCM) Core for Advanced Magnetic Resonance Imaging (CAMRI). Two MRI acquisition protocols were used depending on the clinical trial in which each subject was enrolled. Both protocols included a high resolution T1-weighted magnetization-prepared rapid acquisition gradient-echo (MP-RAGE) sequence (256 sagittal volumes, TR=2.4 s, and flip angle=8°), with protocol-specific differences in echo time (TE=2.41 ms vs 2.24 ms), spatial resolution (voxel size=0.7mm-isotropic vs 0.8mm-isotropic), and field of view (FOV = 224x224x179 mm vs 256x282x205 mm). Diffusion-weighted MRI (dMRI) scans were acquired for all subjects except Cap-DBS3. One dMRI protocol used single-shell, multi-band, high-angular-resolution with 268 axial volumes acquired with anterior-to-posterior phase encoding direction (including 12 b = 0 s/mm^2^ volumes and 256 diffusion-weighted directions at b = 1000 s/mm^2^), along with an additional 7 b = 0 s/mm^2^ axial volumes acquired with posterior-to-anterior phase encoding direction, voxel size=2mm-isotropic, TR=4.2s, TE=65ms, flip angle=90°, FOV=256x256x128 mm, pixel bandwidth=2300 Hz, and imaging frequency around 123.25 MHz. The other protocol used multi-shell, multi-band with 99 axial volumes (including 7 volumes at b=0 s/mm^2^ and 46 diffusion-weighted directions acquired with opposite phase encoding directions at each of two b-values (b=1250 s/mm^2^ and b=2500 s/mm^2^), TR=3.2 s, TE=87 ms, flip angle=90°, voxel size=1.5mm-isotropic, FOV=210x210x138 mm, pixel bandwidth=1700 Hz, and imaging frequency=123.25 MHz. Subjects also underwent intra-operative clinical CT scans obtained post-implantation for lead localizations. CT scans were acquired on a Philips iCT 256 system, with 250-mm reconstruction diameter and contiguous slices of 0.67-mm space between the slices of 0.67 mm thickness. The reconstructed images had an in-plane matrix size of 512 x 512 pixels, and the view size was 1664 x 1236 pixels.

### DBS Programming

DBS was initiated ∼2 weeks post-implantation. Stimulation was optimized through serial programming sessions using clinical assessments of energy, mood, anxiety, and changes in OCD severity between visits ^32^. Clinical response was defined as a ≥35% reduction in Y-BOCS relative to the post-operative, pre-programming baseline ^30^. For subjects Cap-DBS1-3, we extracted the active contacts and stimulation settings from the visit immediately preceding clinical response.

### MRI preprocessing and tractography

#### Preprocessing

Images were first pre-processed using FreeSurfer v7.3.2 of to divide and parcellate brain anatomical structures within each subject’s native space ^33^. Visual inspection was then performed to check for needed corrections to the automatic segmentations.

We preprocessed dMRIs using MRtrix3 tools for denoising ^34^ and Gibbs ring correction ^35^. Next, we performed corrections for susceptibility-induced geometric distortions followed by motion and eddy current correction ^36^. During this step, if diffusion-weighted images were acquired with opposite phase encoding directions, volumes with the same diffusion sensitization were paired and combined to estimate the susceptibility distortion field. If only b = 0 images were available with opposite phase encoding directions, the paired b = 0 images were used for distortion field estimation. Subsequently, we performed bias field correction using Advanced Normalization Tools (ANTs) ^37^. In addition, the skull-stripped b0 image and T1-weighted image were co-registered to generate a FreeSurfer-derived five-tissue-type (5TT) segmentation aligned to the diffusion-weighted images. Skull stripping was then conducted using the FSL’s Brain Extraction Tool ^38 39^.

#### ROI Creation

For each Cap-DBS subject, we created region-of-interest (ROI) masks of their capsulotomies and VTAs in each hemisphere. The capsulotomies were manually segmented from the T1-weighted images by a qualified physician. We created VTA ROIs for the Cap-DBS subjects by localizing the contacts activated at clinical response in MNI space using Lead-DBS v3.0 ^40^ (see Supplementary Methods).

#### Tractography

We performed tractography in three different ways: whole-brain, PFC-constrained, and VTA-seeded.

First, we were interested in any circuits impacted by the capsulotomy and VTA, so we performed whole-brain tractography using Anatomically Constrained Tractography ^41^, adjusting the 5-tissue type (5TT) mask to include streamlines passing through the striatum ^42^. With the “tckgen” function, we generated five million streamlines (maximum angle of 22.5 degrees; maximum length of 250 mm). We then transformed the MICCAI atlas ^43^ into subject-specific space and used the “tckedit” command to determine streamline connectivity.

We were next interested specifically in PFC connections, so we followed a procedure we previously developed to create anatomically plausible PFC streamlines through the ALIC, including in treatment-resistant psychiatric patients ^42,44,45^. An ALIC mask image ROI was extracted from a white matter atlas ^46^, and we subdivided the PFC into 11 subregions per hemisphere using the FreeSurfer’s Desikan-Killiany Atlas ^47^. All ROIs were registered to each subject’s image spaces by applying ANTs-derived transformation matrices. We generated one million streamlines passing through the ALIC and then selected only streamlines passing through relevant ROIs.

### VTA-Seeded Tractography

To investigate subcortical connectivity, we created VTA-seeded tractograms in MRtrix3, with the adjusted 5TT masks. We generated 500,000 streamlines passing through the VTA ROI in each hemisphere. Then, with these streamlines, we used the “tck2connectome” function and the atlas of the Adult Human Hypothalamus (AHH) ^48^ to create connectomes of subcortical connectivity of VTA streamlines.

### Analysis

To quantify and visualize connectivity profiles (Figure 1), we used a custom python script with the “tckinfo” function to extract streamline counts. For the OCD-Control cohort, we averaged the connectivity profiles. Before further processing we removed erroneous regions (e.g., ventricles) and applied a 1% threshold filter. The potentially necessary tracts for successful DBS treatment after failed capsulotomy were calculated by subtracting the capsulotomy connectivity from the joint capsulotomy and VTA connectivity.

Finally, the subcortical connectivity exported from the “tck2connectome” function was visualized as a heatmap in each hemisphere. To understand whether Cap-DBS subcortical connectomes differ from the OCD-Controls, we computed the Pearson’s correlation coefficient between Cap-DBS1&2 and OCD-Controls. Confidence intervals (95%) were estimated using Fisher’s r-to-z transformation and back-transformed to the correlation scale. Scatter plots with a least-squares regression line were generated for visualization. Lastly, we plotted the z-score comparison between Cap-DBS1&2’s connectome to the average OCD-Control connectome created with the Cap-DBS subject’s VTAs.

## 3. Results

### Overview

Although both DBS and capsulotomies targeted the VC/VS region, we found consistent anatomical differences between them. Capsulotomies were located more anteriorly and dorsally, whereas VTAs were positioned posteriorly and ventrally. Capsulotomy ROIs exhibited broad connectivity to frontal cortical and subcortical structures, while VTA ROIs showed comparatively stronger engagement of subcortical pathways. Subtracting capsulotomy connectivity from VTA connectivity identified a small set of potentially necessary tracts dominated by subcortical contributions with inconsistent frontal involvement.

### Locations of VTAs vs capsulotomies

Cap-DBS subjects underwent two invasive procedures (capsulotomy and DBS) that both targeted VC/VS. We observed small but meaningful differences in their placements (Figure 2). VTAs were more ventrally and posteriorly placed than capsulotomies. There were also inter-individual differences: Cap-DBS2 had large bilateral lesions dorsal to the capsulotomies of Cap-DBS1&3.

**Figure 2:**
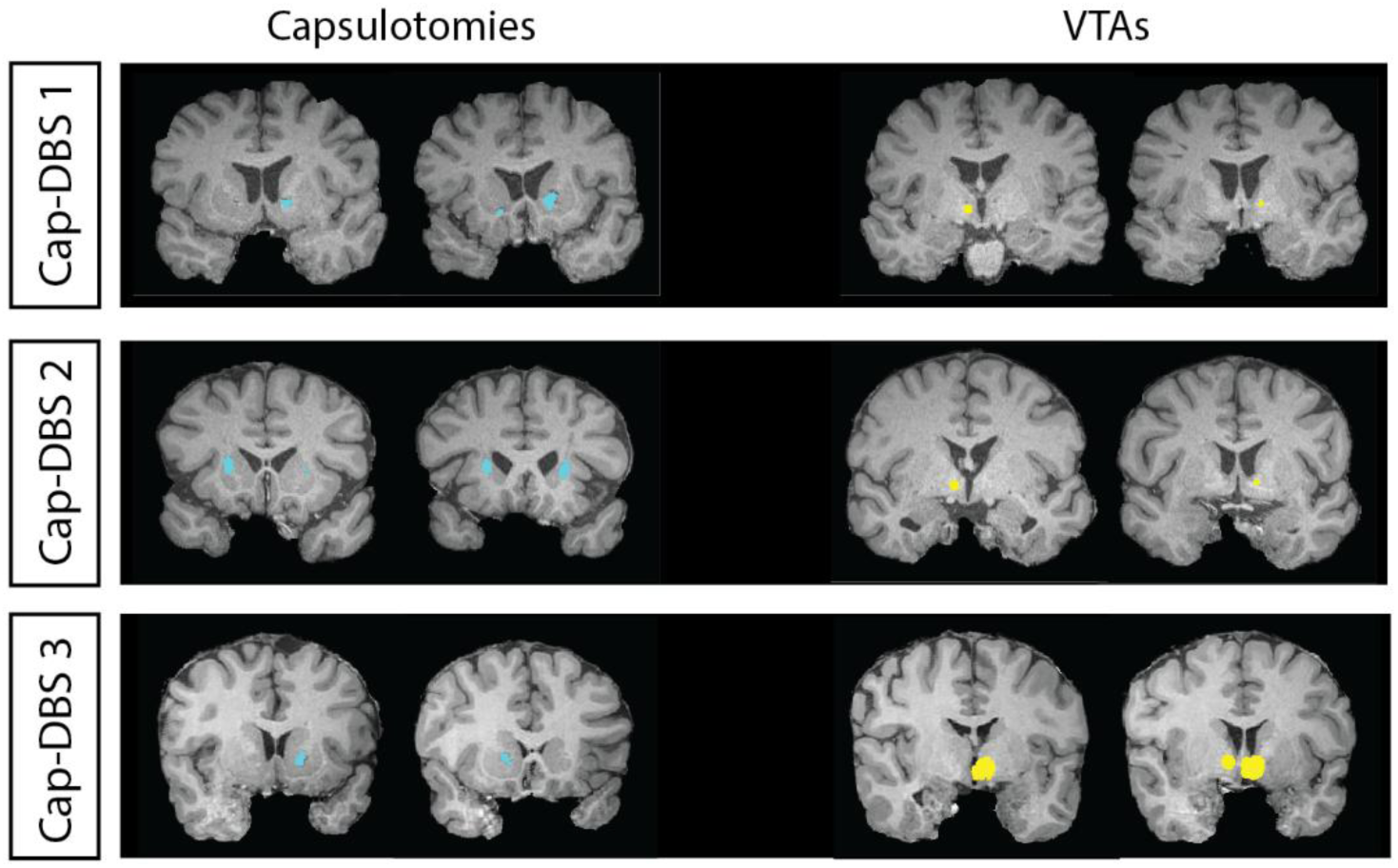
Coronal sections of Cap-DBS subjects. Capsulotomies (blue) and VTAs (yellow) are shown in subject-specific space.

### Capsulotomy connectivity

First, we wanted to understand what connections may have been eliminated by capsulotomy procedure. Because dMRI was not available from prior to capsulotomy, we used two tractography approaches in the OCD-Control cohort to understand the potential connectivity of the capsulotomy region: 1) whole-brain tractography and 2) PFC-constrained tractography.

Overall, all three capsulotomy ROIs displayed extensive connectivity to frontal cortical and subcortical regions (Figure 3). Connectivity of capsulotomy ROIs in OCD-Control whole-brain tractography (Figure 3A) were similar for the ROIs derived from Cap-DBS1&3; Cap-DBS2’s connectivity profile, while containing many of the same regions, had a different distribution. The difference is likely due to the more dorsal placement of the capsulotomy. For all, the left and right hemisphere were similar within-subject. The capsulotomy connectivity profiles in OCD-Control PFC-constrained tractography allowed us to delineate which PFC regions may have lost connections due to the lesion (Figure 3B). Capsulotomy ROIs had the most abundant connections with the superior frontal cortex (SF) and additional connectivity with the central orbitofrontal cortex (COF) and rostral middle frontal cortex (RMF).

**Figure 3:**
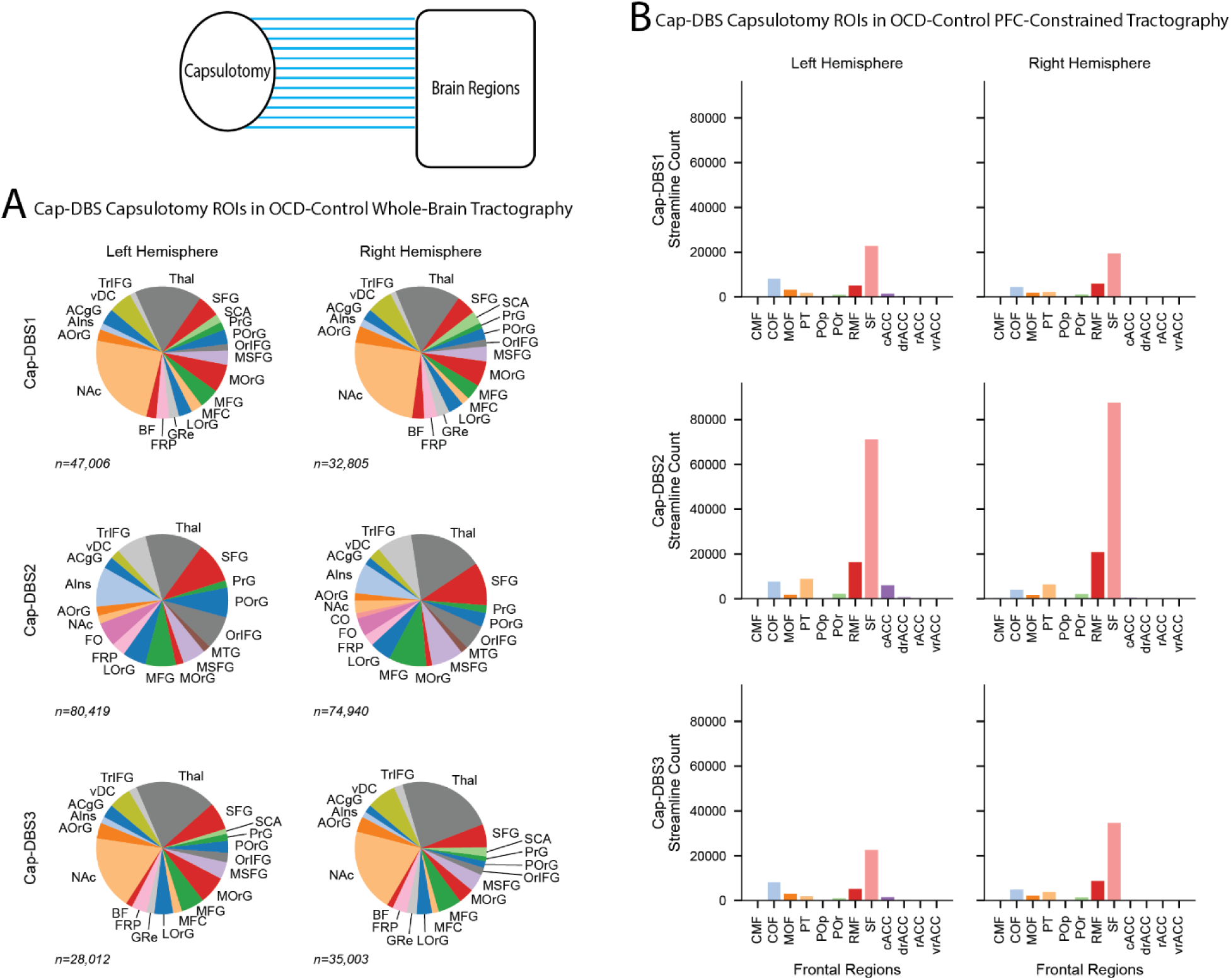
Connectivity profiles of failed capsulotomies. Connectivity profiles of streamlines passing through the capsulotomy ROIs from Cap-DBS subjects in the **A)** OCD-Control cohort whole-brain tractography and **B)** the OCD-Control cohort PFC-constrained Tractography. *Abbreviations (A):* ACgG, Anterior Cingulate Gyrus; AIns, Anterior Insula; AOrG, Anterior Orbital Gyrus; NAc, Nucleus Accumbens Area; Amy, Amygdala; AnG, Angular Gyrus; BF, Basal Forebrain; CO, Central Operculum; Calc, Calcarine Cortex; Cblm, Cerebellum Exterior; Cun, Cuneus; Ent, Entorhinal Area; FO, Frontal Operculum; FRP, Frontal Pole; FuG, Fusiform Gyrus; GRe, Gyrus Rectus; HPC, Hippocampus; IOG, Inferior Occipital Gyrus; ITG, Inferior Temporal Gyrus; LOrG, Lateral Orbital Gyrus; LiG, Lingual Gyrus; MCgG, Middle Cingulate Gyrus; MFC, Medial Frontal Cortex; MFG, Middle Frontal Gyrus; MOG, Middle Occipital Gyrus; MOrG, Medial Orbital Gyrus; MPoG, Postcentral Gyrus Medial Segment; MPrG, Precentral Gyrus Medial Segment; MSFG, Superior Frontal Gyrus Medial Segment; MTG, Middle Temporal Gyrus; OCP, Occipital Pole; OFuG, Occipital Fusiform Gyrus; OpIFG, Opercular Part of the Inferior Frontal Gyrus; OrIFG, Orbital Part of the Inferior Frontal Gyrus; PCgG, Posterior Cingulate Gyrus; PCu, Precuneus; PHG, Parahippocampal Gyrus; PIns, Posterior Insula; PO, Parietal Operculum; POrG, Posterior Orbital Gyrus; PP, Planum Polare; PT, Planum Temporale; PoG, Postcentral Gyrus; PrG, Precentral Gyrus; SCA, Subcallosal Area; SFG, Superior Frontal Gyrus; SMC, Supplementary Motor Cortex; SMG, Supramarginal Gyrus; SOG, Superior Occipital Gyrus; SPL, Superior Parietal Lobule; STG, Superior Temporal Gyrus; TMP, Temporal Pole; TTG, Transverse Temporal Gyrus; Thal, Thalamus Proper; TrIFG, Triangular Part of the Inferior Frontal Gyrus; vDC, Ventral Diencephalon *Abbreviations (B):* CMF, Caudal Middle Frontal Cortex; COF, Central Orbitofrontal Cortex; MOF, Medial Orbitofrontal Cortex; PT, Pars Triangularis; POp, Pars Opercularis; POr, Pars Orbitalis; RMF, Rostral Middle Frontal Cortex; SF, Superior Frontal Cortex; cACC, Caudal Anterior Cingulate Cortex; drACC, Dorsal Rostral Anterior Cingulate Cortex; rACC, Rostral Anterior Cingulate Cortex; vrACC, Ventral Rostral Anterior Cingulate Cortex.

### VTA connectivity

We asked what regions may have been modulated by the DBS if performed in brains without lesions (with VTAs derived from Cap-DBS patients). In the OCD-Control cohort, there was some connectivity of the VTA with frontal regions, but VTA ROIs had a preponderance of connections with the thalamus and ventral diencephalon (vDC). The PFC-constrained tractography (Figure 4B) showed a higher number of streamlines in the right hemisphere than the left. In the right hemisphere, the RMF, SF, COF and medial orbitofrontal cortex (MOF) stood out as having high streamline counts. In the left hemisphere, for all VTAs, streamline counts were low.

**Figure 4:**
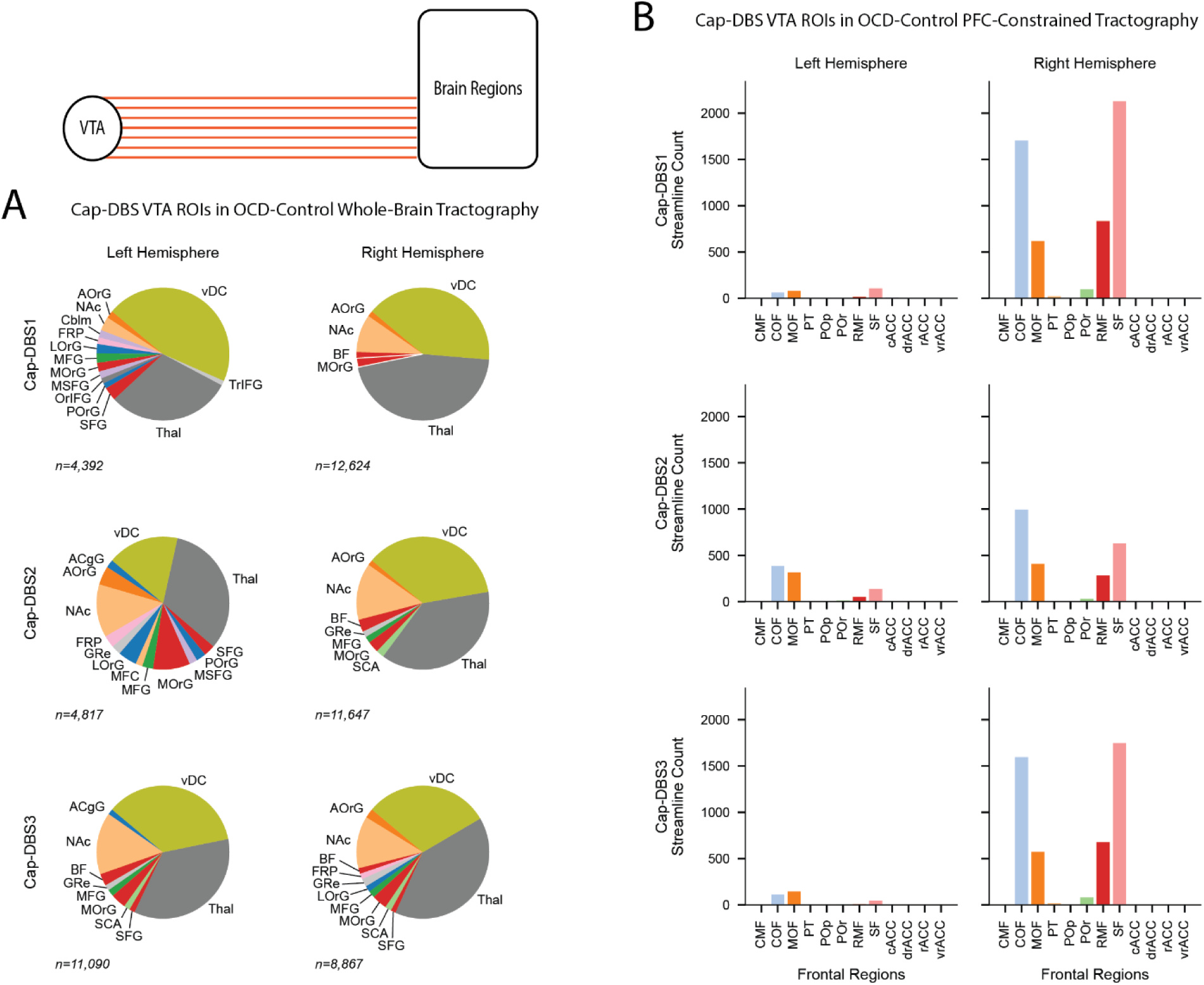
Connectivity profiles of (effective) VTAs. Connectivity profiles of streamlines passing through the VTA ROIs from Cap-DBS subjects in the **A)** OCD-Control cohort whole-brain tractography and **B)** the OCD-Control cohort PFC-constrained Tractography. Abbreviations as in Figure 3.

### Potentially necessary tracts

By subtracting the capsulotomy streamlines from the VTA streamlines, we identified connections that would have been impacted by DBS treatment but not eliminated by the lesion. These “potentially necessary tracts” represent which connections may have been part of the therapeutic mechanism in the Cap-DBS patients (with the understanding that lesioned tracts may also be necessary, but not sufficient, for improvement). In OCD-Control whole-brain tractography (Figure 5A), the thalamus, vDC, and nucleus accumbens area (NAc) made up a large proportion of the connections of the potentially necessary tracts.

**Figure 5:**
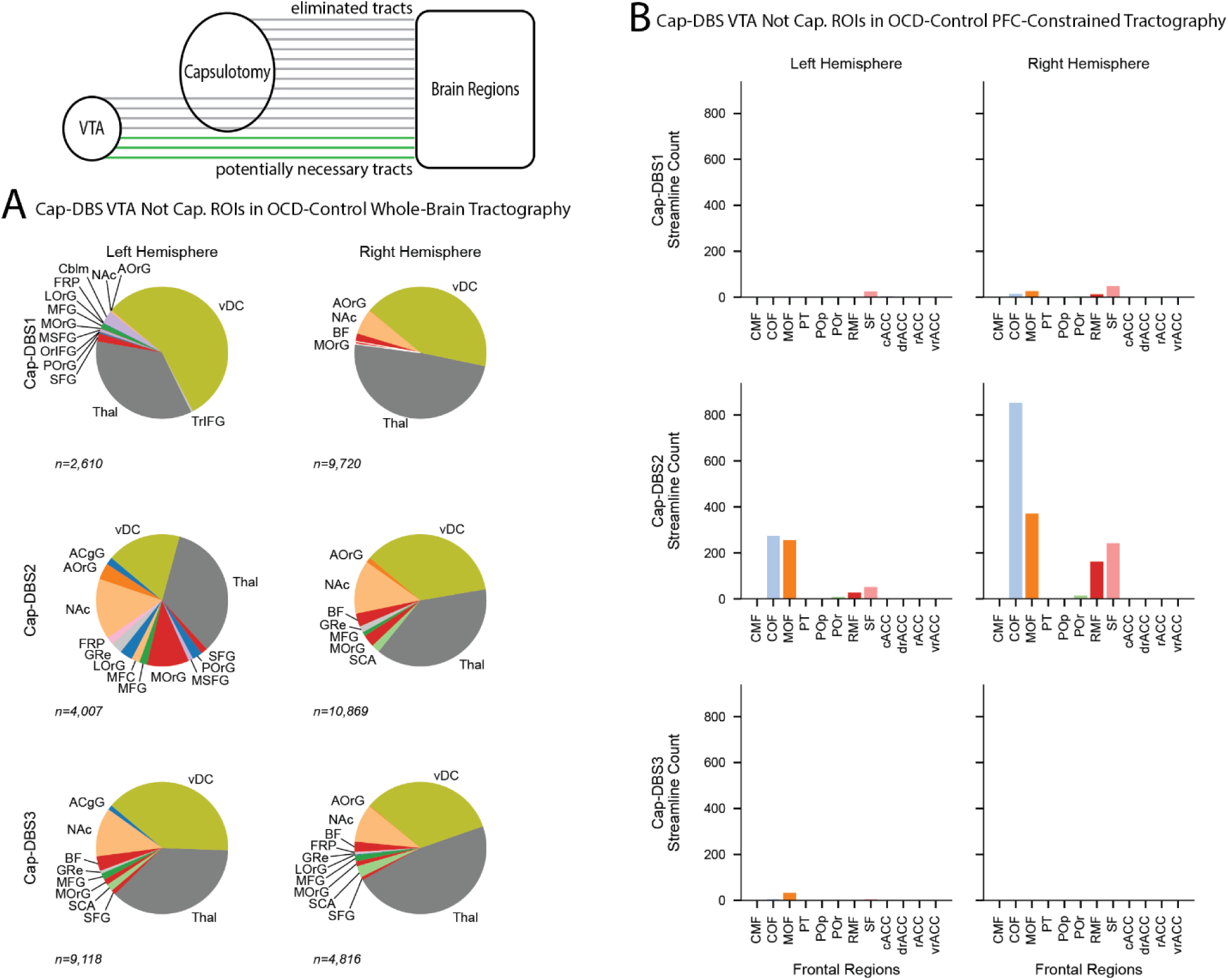
Connectivity of potentially necessary tracts. Connectivity profiles of streamlines passing through the VTA but *not* capsulotomy ROIs from Cap-DBS subjects in the **A)** OCD-Control cohort whole-brain tractography and **B)** the OCD-Control cohort PFC-constrained tractography. Abbreviations as in Figure 3.

In the PFC-constrained tractography in OCD-Controls (Figure 5B), profiles from Cap-DBS1&3 had very few potentially necessary streamlines to frontal cortical regions. Connectivity profiles from Cap-DBS2 had more frontal connectivity of potentially necessary tracts. In both hemispheres, the largest connections were to the COF, MOF, RMF, and SF. Notably, even in cases where PFC fibers were present in this analysis, they were inconsistent across hemispheres and subjects.

In tractography performed specifically in Cap-DBS1&2 before DBS but after capsulotomy (Figure 6), potentially necessary tracts connected the most to the thalamus, NAc, and vDC. The connectivity in Cap-DBS2’s left hemisphere included more cortical regions (Figure 6B). Results from PFC-constrained tractography (Figure 6C) were markedly different between Cap-DBS1&2. Potentially necessary tracts from Cap-DBS1 were fewer than that of Cap-DBS2. Tracts from Cap-DBS1 had the most connectivity to the SF followed by RMF. Cap-DBS2 had more potentially necessary tracts, particularly in the right hemisphere. The largest connections in descending order for both hemispheres were COF, MOF, RMF, and SF.

**Figure 6:**
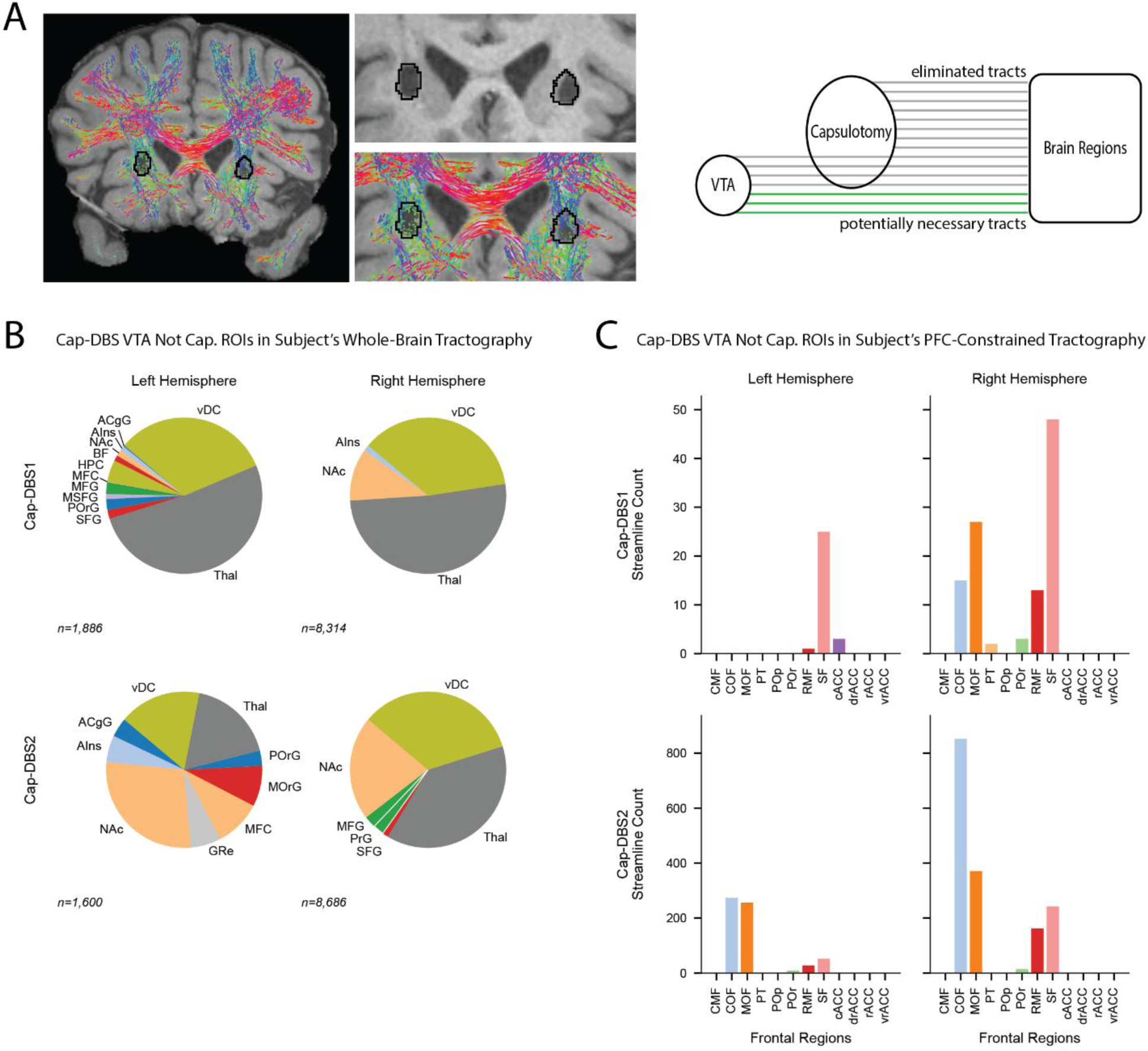
Connectivity of potentially necessary tracts in subject-specific whole-brain tractography. **A)** Streamlines are much less prominent in the capsulotomy ROI (outlines). **B)** Connectivity profiles of potentially necessary tracts in Cap-DBS subject-specific whole-brain tractography. **C)** Connectivity to frontal regions of potentially necessary tracts using Cap-DBS subject-specific PFC-constrained tractography. Abbreviations as in Figure 3.

### Intra-subcortical connectivity

We next wanted to investigate the involvement of subcortical regions in more detail. In the connectome from Cap-DBS1’s left hemisphere in subject specific VTA-seeded tractography (Figure 7A) there were connections to the inferior thalamic peduncle (ITP); the largest edges were with the substantia nigra (SN) and red nucleus (RN). In the right hemisphere the largest connections were the nucleus basalis of Meynert (NBM) with the fornix (Fx) and ITP. The AHH connectome from Cap-DBS2 had more strong connections than Cap-DBS1’s. In the left hemisphere BNST was moderately connected to the lateral hypothalamus (LH), mammillothalamic tract (MTT), and RN. The diagonal band of Broca (DBB) was moderately connected to the ITP. The strongest connectome edge in Cap-DBS2’s connectome was between the Fx and NBM. In the right hemisphere the strongest connections were between the NBM and the DBB, Fx, and LH. There were moderate connections of the DBB, Fx, and LH with the ITP.

**Figure 7:**
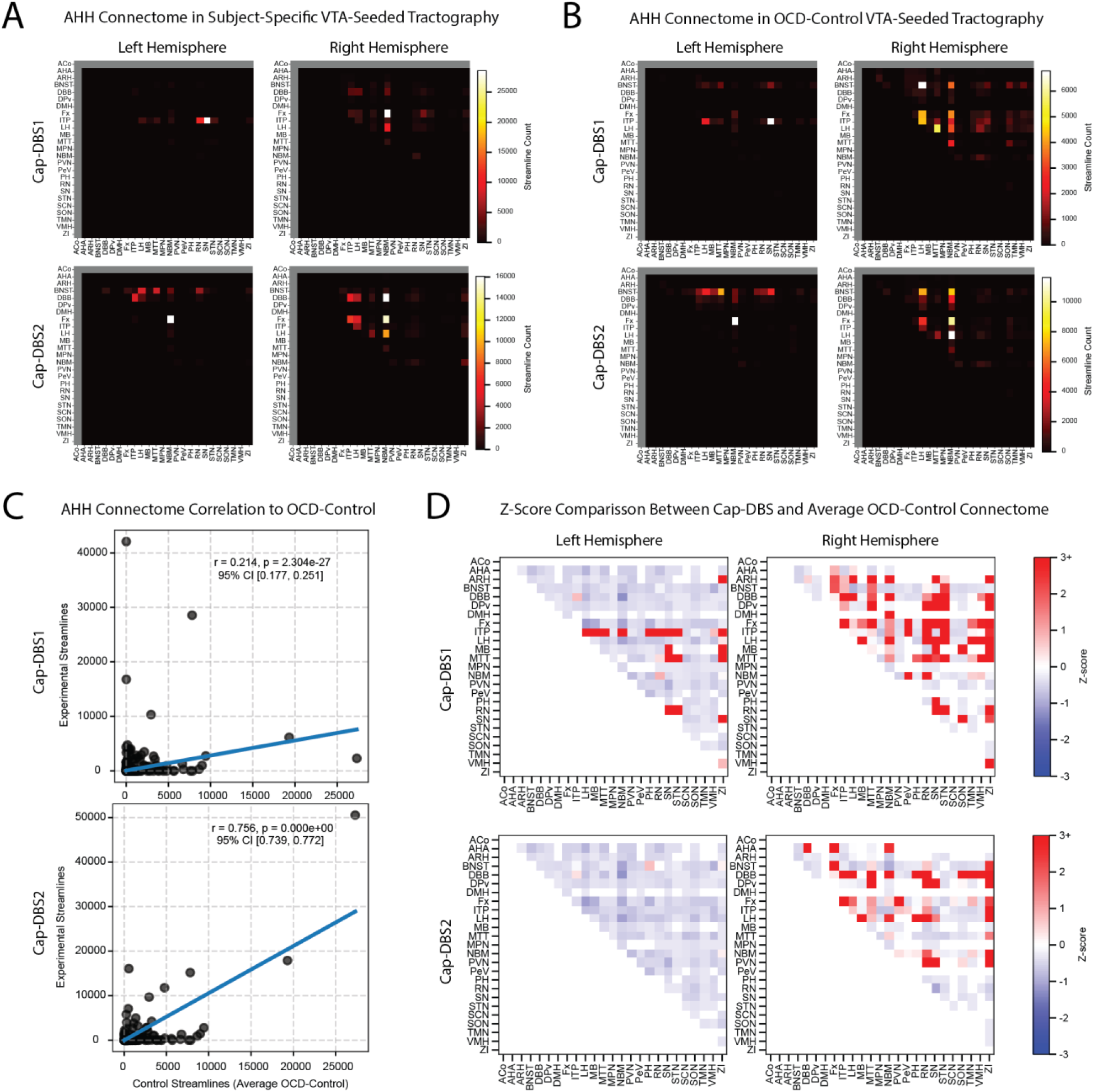
Intra-subcortical connectomes. **A)** Subcortical (AHH) connectome of Cap-DBS subject-specific VTA-seeded tractography. **B)** Subcortical (AHH) connectome of the averaged OCD-Control VTA-seeded tractography. **C)** Correlation of subcortical connectomes from Cap-DBS1&2, respectively, to the average OCD-Control connectomes. **D)** Z-scores of connectome differences between Cap-DBS1&2, respectively, to the average OCD-Control connectomes. Red indicates stronger connections in the cap-DBS subject’s connectome. ACo, Anterior Commissure; AHA, Anterior Hypothalamic Area; ARH, Arcuate Hypothalamic Nucleus; BNST, Bed Nucleus of the Stria Terminalis; DBB, Diagonal Band of Broca; DPv, Dorsal Periventricular Hypothalamic Nucleus; DMH, Dorsomedial Hypothalamic Nucleus; Fx, Fornix; ITP, Inferior Thalamic Peduncle; LH, Lateral Hypothalamus; MB, Mammillary Bodies; MTT, Mammillothalamic Tract; MPN, Medial Preoptic Nucleus; NBM, Nucleus Basalis of Meynert; PVN, Paraventricular Nucleus; PeV, Periventricular Hypothalamic Nucleus; PH, Posterior Hypothalamic Nucleus; RN, Red Nucleus; SN, Substantia Nigra; STN, Subthalamic Nucleus; SCN, Suprachiasmatic Hypothalamic Nucleus; SON, Supraoptic Hypothalamic Nucleus; TMN, Tuberomammillary Hypothalamic Nucleus; VMH, Ventromedial Hypothalamus; ZI, Zona Incerta.

The averaged OCD-Control AHH connectome (Figure 7B) revealed a similar pattern of hypothalamic and basal forebrain connectivity, particularly involving the BNST, LH, and NBM. However, the relative strengths and distribution of these edges differed between the Cap-DBS subjects and the control average, suggesting subject-specific deviations in hypothalamic circuitry.

To quantify similarity between Cap-DBS and control connectivity profiles, we correlated the AHH connectomes from each Cap-DBS subject with the averaged OCD-Control connectome (Figure 7C). Cap-DBS1’s connectivity profile demonstrated a weak but significant correlation with the control connectome (r = 0.214, p = 2.304 × 10^⁻²⁷^, 95% CI [0.177, 0.251]). In contrast, Cap-DBS2’s connectivity profile showed a substantially stronger correspondence with the control connectome (r = 0.756, p ≈ 0, 95% CI [0.739, 0.772]), suggesting that the overall pattern of VTA–subcortical connectivity in Cap-DBS2 more closely resembles the control cohort.

Finally, Z-score comparisons were computed to identify connections that deviated from the control distribution (Figure 7D). Across both Cap-DBS subjects, deviations were more prominent in the right hemisphere. Cap-DBS1 exhibited several connections with increased strength relative to controls (positive Z-scores), while many other edges were weaker than the control mean. Cap-DBS2 showed a similar pattern but with fewer large deviations overall, consistent with the stronger global correlation observed in the correlation (Figure 7C). Notably, several right-hemisphere connections involving the DBB, LH, NBM, and associated hypothalamic pathways displayed positive Z-scores, indicating stronger connectivity relative to the OCD-Control cohort.

## 4. Discussion

We had the unique opportunity to study connectivity profiles from patients that had unsuccessful capsulotomy followed by successful DBS treatment. Our multi-pronged tractography analysis of the connectivity profiles of capsulotomy, VTA, and potentially necessary tracts revealed a surprising lack of consistent PFC involvement in potentially necessary tracts for successful DBS. Capsulotomy ROIs involved PFC and subcortical regions, while VTA and potentially necessary tracts mostly involved subcortical regions, even when special care was taken to preserve the anatomy of connections to the PFC. Further investigation of subcortical connections showed that, although the connectivity of the Cap-DBS subjects’ subcortical regions is similar to that of the OCD control group, there are several regions that have relatively stronger connections. These differences may be due to remodeling after the capsulotomy. Overall, the surprising dominance of subcortical connections in the connectivity profiles of VTAs and potentially necessary tracts points to a potentially stronger role of intra-subcortical connectivity DBS treatment.

Neurosurgical approaches to OCD treatment have been available to patients since the mid-twentieth century ^49^, but the location of the interventions has shifted over time. The first gamma knife capsulotomy was placed in the middle of the ALIC. Future attempts have covered the ventral ALIC. Voxel-wise lesion-symptom mapping has suggested that more posterior and dorsal lesion sites within the ventral ALIC are related to greater Y-BOCS reduction ^50^. Early applications of DBS for OCD targeted similar locations to the capsulotomy ^21^. Over time, DBS targets have moved more posteriorly within the ALIC toward the BNST ^23,24^, corresponding with improved DBS outcomes^22^. This posterior location may have a denser convergence of fibers connected to distinct brain regions ^51^. However, the effectiveness of capsulotomies and DBS for tr-OCD remains comparable.

Of course, stimulation and lesions are not equivalent methods, even when placed in the same location. The physiological effect of each intervention may be different, and thus the connections that are necessary to modify may be accordingly different. The mechanism of DBS is still unclear. Initially DBS was thought to serve as a functional lesion that could be turned on and off ^52–54^. The current understanding of DBS is that several mechanisms can be at play: local inhibition, local excitation, modulation of temporal coding, and enhancement of neurogenesis ^52^.

In a large longitudinal analysis that included post-treatment MRI, Chen et al.^55^ compared the circuit mechanisms of capsulotomy and DBS treatments. They identified both shared and distinct network involvement of the two treatment types. In particular, both DBS and capsulotomy led to enhanced cortico-cortical but reduced frontal cortico-subcortical functional connectivity, and these changes were linked with improved clinical outcomes.

These results strongly suggest that the PFC-subcortical connections that were disrupted in the failed capsulotomies in our patients were a necessary part of treatment. Considering their results in combination with our own, disrupting frontal cortico-subcortical connections may drive therapy by weakening cortico-subcortical connectivity and enhancing cortico-cortical and intra-subcortical connections. Such a mechanism may explain the enhanced connectivity of subcortical regions in our Cap-DBS subjects’ subcortical connectomes when compared to the OCD control group. Indeed, a PET study comparing the brain effects of capsulotomy to DBS found, consistent with our findings, wider spread and more subcortical changes with capsulotomy relative to DBS ^56^.

## 5. Conclusion

This study leverages a rare and clinically informative patient population to advance understanding of the circuit mechanisms underlying effective neuromodulation for tr-OCD. Still, several limitations should be acknowledged. First, the sample size is necessarily small, reflecting the rarity of patients who undergo both capsulotomy and DBS, and limits the generalizability of the results. Second, dMRI was unavailable prior to capsulotomy, requiring reliance on control cohorts and modeling approaches to infer pre-lesion connectivity. Third, tractography-based methods are inherently limited in their ability to infer directionality, synaptic strength, or causal influence, and should be interpreted as probabilistic representations of anatomical pathways rather than definitive circuit maps. Finally, the absence of analyzable diffusion data in one subject constrained subject-specific comparisons. Despite these limitations, the consistency of findings across analytic approaches and cohorts strengthens the central conclusions.

Importantly, this work contributes to a shift in the field away from viewing OCD solely as a disorder of PFC-subcortical dysregulation and toward a broader network perspective that includes intra-subcortical systems. By highlighting subcortical connectivity as a prominent feature of effective DBS following failed capsulotomy, this study helps refine hypotheses about the mechanisms of neuromodulation and underscores the need to consider intra-subcortical interactions in both therapeutic targeting and mechanistic models. These insights may inform future efforts to personalize neurosurgical treatments, guide electrode placement, and integrate connectivity-based biomarkers into predictive frameworks. Ultimately, continued integration of detailed circuit mapping, longitudinal clinical outcomes, and individualized imaging may optimize neuromodulation strategies for those with tr-OCD.

## Data Availability

All data produced in the present study are available upon reasonable request to the authors.

## Acknowledgments

We acknowledge funding from NIH grants R01MH139889 (NRP), UM1NS32207 (SRH), UH3NS100549 (WKG), and the Robert and Janice McNair Foundation (NRP, SRH, SAS).

