## Supplemental Methods for "Neural basis of successful DBS for OCD after failed capsulotomy"

**Supplementary Materials**

**Supplementary Methods**

*Participant response timelines.* Cap-DBS1 had three capsulotomy procedures; he was between 25 and 34 years old at the time of his two Gamma Knife capsulotomies and was between 35 and 44 years old at the time of his stereotaxic laser ablation capsulotomy. After no improvement (Y-BOCS score of 40 post-capsulotomy), he underwent DBS 2.6 years after the capsulotomy. Following DBS, he had a Y-BOCS score of 25 (37% reduction from baseline). Cap-DBS2 was between 25 and 34 years old at the time of radiosurgical anterior capsulotomy, but his symptoms worsened to a Y-BOCS score of 34. He then underwent DBS that resulted in a Y-BOCS score of 18 (47% reduction from baseline). Cap-DBS3 was between 45 and 54 years old at the time of radiosurgical capsulotomy. After no improvement (Y-BOCS score of 30), he underwent DBS. His Y-BOCS at follow up from DBS was 19 (37% reduction from baseline). The current paper used data from two clinical trials (NCT04806516 and NCT05915741).

*VTA calculation.* We began with a linear coregistration of the patient’s preoperative MRI and postoperative CT with Advanced Normalized Tools (ANTs) ^1^. Coregistered images were then normalized to the MNI non-linear 2009b space using ANTs Symmetric Normalization (SyN) algorithm and brainshift- corrected using the coarse mask tool to account for postoperative linear deformation of brain tissue ^2^. Following coregistration, electrode trajectories were automatically pre-reconstructed using the Precise and Convenient Electrode Reconstruction for Deep Brain Stimulation (PaCER) toolbox, with manual refinements as needed ^3^. VTAs were created with the SimBio/FieldTrip model ^4^ using the stimulation parameters that immediately preceded clinical response. The binary ROI masks in MNI space for each hemisphere were exported to use in tractography analysis.
